# A NATUrally Randomized trial Evaluating a vaccine-like strategy to lower LDL by inhibiting PCSK9 on the lifetime risk of major cardiovascular events (NATURE-PCSK9)

**DOI:** 10.1101/2024.06.30.24309740

**Authors:** Brian A Ference, Thatcher B. Ference, Alberico L Catapano, Stephen J. Nicholls, Kausik K Ray

**Affiliations:** DeepCausalAI Institute for Clinical Translation, Cambridge, U.K.; Department of Pharmacological and Biomolecular Sciences, University of Milan and Multimedica IRCCS, Milano, Italy; Victorian Heart Institute, Monash University, Melbourne, Australia; Imperial Centre for Cardiovascular Disease Prevention, Department of Primary Care and Public Health, School of Public Health, Imperial College London, London U.K

## Abstract

**Background:** Maintaining low levels of low-density lipoproteins (LDL) over time has the potential to substantially reduce the lifetime risk of atherosclerotic cardiovascular disease. However, the optimal timing of lowering LDL to prevent atherosclerotic cardiovascular events is unknown.

**Methods:** We combined evidence from Mendelian randomization studies and randomized trials to develop a causal AI algorithm to estimate the benefit of lowering LDL on the risk of major cardiovascular events (MCVE) in discrete time-units of exposure. We tested the accuracy of this algorithm among 440,371 participants randomized by nature to a partial loss-of-function (LOF) variant in the PCSK9 gene, and 46,488 participants in two large randomized trials of PCSK9 inhibitors. We then used this algorithm to estimate the benefit of lowering LDL using a once-yearly dose of an siRNA directed against PCSK9 beginning at different ages among 2.3 million men and women.

**Results:** The causal AI algorithm accurately estimated the benefit of lifelong lower LDL due to partial loss-of-function of the PCSK9 gene, and the benefit of lowering LDL with a PCSK9 inhibitor starting at a mean age of 61 years, with nearly superimposable observed and predicted event curves. Lowering LDL by 36% was estimated to reduce the lifetime risk of MCVE by 57% (HR: 0.43, 95%CI: 0.39-0.47) if started at age 30, by 48% (HR: 0.52, 95%CI: 0.50-0.54) if started at age 40, by 38% (HR: 0.62, 95%CI: 0.60-0.65) if started at age 50, and by 26% (HR: 0.74, 95%CI: 0.71-0.77) if started at age 60 years. Moderate LDL lowering starting at age 40 years was estimated to have a greater benefit than more aggressively LDL lowering beginning at age 55 years, with a lower residual risk at all ages. In addition, the benefit of earlier LDL lowering persisted throughout life leading to a quantifiable legacy benefit.

**Conclusions:** The benefit of lowering LDL is determined by the magnitude, duration, and timing of LDL lowering. Modest sustained LDL lowering beginning in early to middle adulthood, which can be achieved with a once-yearly dose of a PCSK9 siRNA, may be the optimal strategy to prevent atherosclerotic cardiovascular events by slowing the progression of atherosclerosis.

## INTRODUCTION

Atherosclerosis is caused by the progressive trapping of low density lipoproteins (LDL) and other apolipoprotein-B100 containing lipoproteins within the arterial wall.^1,2,3^ As more lipoproteins become trapped within the artery wall over time, the atherosclerotic plaque gradually enlarges and the risk of having an acute cardiovascular event increases directly proportional to the size of the accumulated plaque burden.^4,5^ Therefore, maintaining low levels of LDL beginning early in life could potentially substantially reduce the lifetime risk of cardiovascular events by slowing the progression of atherosclerosis.^2,4^

Indeed, Mendelian randomization studies suggest that lifelong exposure to lower LDL is associated with a much greater reduction in the risk of cardiovascular events as compared to the same magnitude of LDL lowering started later in life in randomized trials.^6,7,8^ Recently, a small interfering RNA (siRNA) therapy directed against PCSK9 was shown to reduce plasma LDL cholesterol (LDL-C) by a time-averaged 36% with a single yearly dose.^9^ The durability of this LDL lowering effect suggests that this therapy could potentially be used in an annual ‘vaccine-like’ strategy to reduce the lifetime risk of atherosclerotic cardiovascular events by maintaining lower LDL levels to slow the progression of atherosclerosis.^10^

However, Mendelian randomization studies only estimate the benefit of maintaining lower LDL throughout life beginning at birth, while randomized trials only estimate benefit of lowering LDL for a few years starting late in life.^11^ Therefore, the benefit of lowering LDL at all ages between these two extremes is unknown. The objective of this study was two-fold. First, to encode the biological effect of LDL on the risk of atherosclerotic cardiovascular events into a causal AI algorithm; and second, to use this algorithm to estimate the potential benefit of lowering LDL using an annual dose of an siRNA directed against PCSK9 beginning at different ages. The overall goal was to make inferences about the optimal timing and intensity of lowering LDL to prevent atherosclerotic cardiovascular events by slowing the progression of atherosclerosis.

## METHODS

### STUDY DESIGN

The study proceeded in several steps. First, we developed a causal AI algorithm to estimate the benefit of lowering LDL on the risk of cardiovascular events beginning at any age and extending for any duration according to the protocol described in the Supplement. Briefly, we calculated the instantaneous hazard ratio per unit lower LDL by comparing the observed event rates and differences in LDL levels during every year of life among participants randomized by nature to one or more genetic variants associated with lower LDL through the LDL receptor pathway (Supplemental Table 1). In addition, we calculated the instantaneous hazard ratio per unit lower LDL by comparing the observed event rates and differences in LDL levels during every month of follow-up among participants randomized to treatment with a statin or placebo in randomized cardiovascular outcomes trials (Supplemental Table 2). We then combined this unconfounded randomized evidence to train an ensemble of deep and machine learning algorithms to learn a function for estimating the causal effect of lower LDL on the risk of cardiovascular events in discrete time units of exposure, conditional on previous exposure to take into account the disease burden that accumulates prior to LDL lowering. Second, we evaluated the accuracy of the causal AI algorithm for estimating the benefit of lowering LDL by comparing the observed and predicted event curves during every year of life among participants randomized by nature to lower LDL due to a partial loss-of-function (LOF) variant in the PCSK9 gene (that was not included in the derivation stage); and the observed and predicted event curves among participants randomized to treatment to lower LDL with a monoclonal antibody (mAb) directed against PCSK9 or placebo in two large randomized cardiovascular outcome trials.^12,13^

Third, we then used the causal AI algorithm to compare the estimated benefit of lowering LDL by 36%, or approximately what can be achieved with a once-yearly dose of an siRNA directed against PCSK9, beginning at ages 30, 40, 50, and 60 years on the lifetime risk of cardiovascular events among 1.1 million men and 1.2 million women enrolled the UK General Practice Research Database.^14^ Fourth, we used the causal AI algorithm to compare the estimated benefit of lowering LDL by 36% beginning at 40 years with the benefit of lowering LDL by 52%, or approximately what can be achieved with twice-yearly doses of a PCSK9 siRNA,^15,16^ beginning at age 55 years to make inferences about the relative contribution of the magnitude and duration of LDL lowering on lifetime risk. Fifth, we used the causal AI algorithm to compare the estimated benefit of lowering LDL by 36% beginning at ages 30, 40, and 55 years on the remaining lifetime risk of cardiovascular events among persons who survive to age 55 without a cardiovascular event, to quantify the potential legacy benefit of earlier LDL lowering.^17^

### STUDY POPULATION

The causal AI LDL lowering benefit algorithm was derived using individual participant data from 443,641 participants enrolled in the UK Biobank who self-identified as being of European ancestry, and did not have any documented hospital, general practice or self-reported diagnosis of myocardial infarction, coronary revascularization, stroke (of any type), transient ischemic attack, angina, diabetes, or cancer before the age of 30 years.^18^ This data was supplemented with summary level data from up to 22 randomized statin trials enrolling a total of 142,231 participants.^19,20^ The reported event curves from each trial were used to construct a lifetable to recover the instantaneous hazard of experiencing the selected outcome during each interval of follow-up among participants who had not experienced an outcome event at the start of that interval. The instantaneous hazard ratio (HR) during a selected interval of follow-up was estimated as the ratio of the interval hazards of experiencing a cardiovascular event in either treatment group. The standard error (SE) was estimated from the reported number of participants who remained at risk at the beginning of each interval (which excludes participants who either experienced a major cardiovascular event or were censored prior to the start of that follow-up interval).

The accuracy of the causal AI algorithm for predicting long-term benefit of lowering LDL was evaluated among the 440,371 UK Biobank participants who were randomized by nature to a partial LOF variant in PCSK9 gene (which was not included in the training data). The accuracy for predicting short-term benefit of lowering LDL was evaluated among the 46,488 participants in the FOURIER and ODYSSEY Outcomes trials who were randomized to a mAb directed against PCSK9.^12,13,21^ The causal AI algorithm estimated benefit of lowering LDL beginning at different ages and dosing schedules was evaluated among 2.3 million participants from the UK General Practice Research Database for whom the composite rate of incident cardiovascular events and death was available during every month of life between the ages of 30 and 90 years.^13^ This information was used to construct separate lifetables among men and women for analyses adjusted for the competing risk of death.

### STUDY EXPOSURES AND OUTCOMES

The primary exposure was difference in plasma LDL levels between the groups being compared. LDL was measured at the time of enrolment and compared at each age among participants randomized by nature to one or more variants associated with lower LDL among participants in the UK Biobank; and measured at the time of enrolment and compared during each follow-up interval among participants enrolled in the randomized trials. The primary outcome was major cardiovascular events (MCVE), defined as the first occurrence of either a fatal or non-fatal myocardial infarction, fatal or non-fatal ischemic stroke, or coronary revascularization.

### STATISTICAL ANALYSIS

The primary analysis used sex-specific Cox proportional hazards models in time-to-event analyses with age as the time scale. The date of all events was recorded from hospital episode statistics, general practice records, or self-reported. Each participant was censored at the age they experienced either a primary outcome event, death due to a cause other than MI or ischemic stroke (treated as a competing risk), or the age at last reported follow-up. A 2-tailed P value less than 0.05 was considered statistically significant. All analyses were performed using Stata (version 18; StataCorp), R (version 3.3.3), or Python (version 3.7). Further details are provided in the supplement methods.

## RESULTS

### STUDY POPULATION

A description of the 443,641 UK Biobank participants who contributed individual data to derive the casual AI LDL lowering benefit algorithm is provided in Supplemental Table 3. A total of 36,205 of these participants experienced a first major cardiovascular event between the ages of 30 and 80 years during 16,986,996.6 person-years of follow-up.

Among the 440,371 UK Biobank participants with genotype data available for rs11591147, 54% were female, the median age at enrolment was 58.6 years, the median age at censoring was 69.2 years. There were no differences in baseline characteristics among 15,341 (3.5%) PCSK9 partial LOF variant carriers compared to the 425,030 non-carriers (except for lipid and lipoprotein levels) thus demonstrating that allocation was approximately random (Supplemental Table 4). The median time-averaged difference in plasma LDL-C levels between partial LOF carriers and non-carriers was 12.0 mg/dL (8.8%) among all participants, and 14.5 mg/dL (10.2%) among participants not on lipid lowering therapy at the time of enrolment. These differences varied modestly by age (Supplemental Figure 1).

### COMPARING OBSERVED AND PREDICTED BENEFIT OF LOWER LDL

A total of 35,806 participants experienced a first major cardiovascular event between the ages of 30 and 80 years during 16,780,207.2 person-years of follow-up. Participants with a partial LOF variant in the PCSK9 gene had a 24% lower total lifetime risk of major cardiovascular events (HR: 0.76, 95%CI: 0.71-0.82, p=7.5×10^-13^). The causal AI algorithm accurately estimated the benefit of lifelong lower LDL due to partial LOF of the PCSK9 gene for the observed differences in LDL between the groups at all ages, with nearly superimposable predicted and observed event curves (Figure 1A).

**Figure 1:**
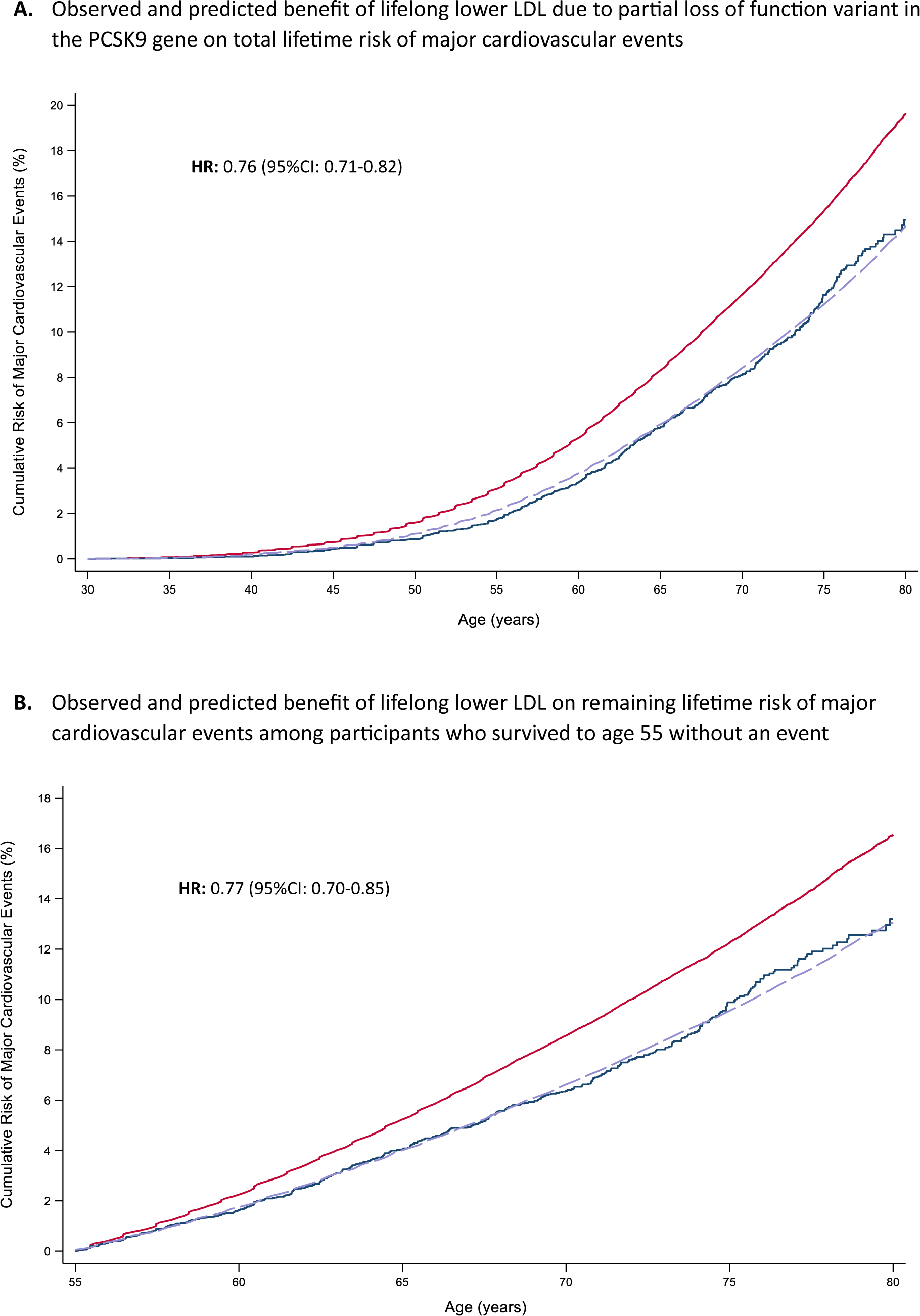
Observed and Predicted Benefit of Lifelong LDL Lowering on Lifetime Risk of Major Cardiovascular Events. Panel A shows the cumulative rate of major cardiovascular events during 16,780,207 person-years of follow-up among 440,371 participants enrolled in the UK Biobank among whom 35,806 participants experienced a first major cardiovascular event between the ages of 30 and 80 years. The red line shows the cumulative event curve for the 425,030 participants without a partial loss-of-function variant in the PCSK9 gene, and the blue line shows the cumulative event rate among the 15,341 (3.5%) participants who were randomized by nature to a PCSK9 partial loss-of-function LOF variant resulting in lifetime exposure to lower LDL. The dashed lavender line represents the cumulative event at all ages predicted by the causal AI algorithm for participants with a partial LOF variants based on the observed differences in LDL between the two groups over time. Panel B shows the cumulative event curves during 16,780,207 person-years of follow-up among participants who survived to age 55 without a major cardiovascular event. The dashed lavender line represents the cumulative event curve predicted by the causal AI algorithm for participants with a partial LOF variants who survived to age 55 without a event. HR is hazard ratio, and CI is confidence interval.

Similarly, among participants who survived to age 55 without a MCVE, a total of 21,561 participants experienced a first MCVE between the ages of 55 and 80 years during 5,931,657 persons-years of follow-up. Participants with a partial LOF variant in the PCSK9 gene had a 23% lower remaining lifetime risk of major cardiovascular events (HR: 0.77, 95%CI: 0.70-0.85, p=1.9×10^-7^). Once again, the causal AI algorithm accurately estimated the benefit of lifelong lower LDL due to partial LOF of the PCSK9 gene for the observed differences in LDL between the groups at all remaining ages between 55 and 80 years, with nearly superimposable predicted and observed event curves (Figure 1B).

Among the 27,564 participants enrolled in the FOURIER trial, 1,829 participants experienced a first major cardiovascular event during a median of 2.2 years of follow-up.^11,20^ Participants randomized to treatment with a PCSK9 mAb beginning at an average age of 62.5 years had a time-averaged 53.4 mg/dL lower LDL and a 20% lower risk of major cardiovascular events (HR: 0.80, 95%CI: 0.73-0.88, p=1.2×10^-8^). Once again, the causal AI algorithm accurately estimated the benefit of lowering due to treatment with a PCSK9 mAb during every month of follow-up during the trial, with nearly superimposable predicted and observed event curves (Figure 2A).

**Figure 2:**
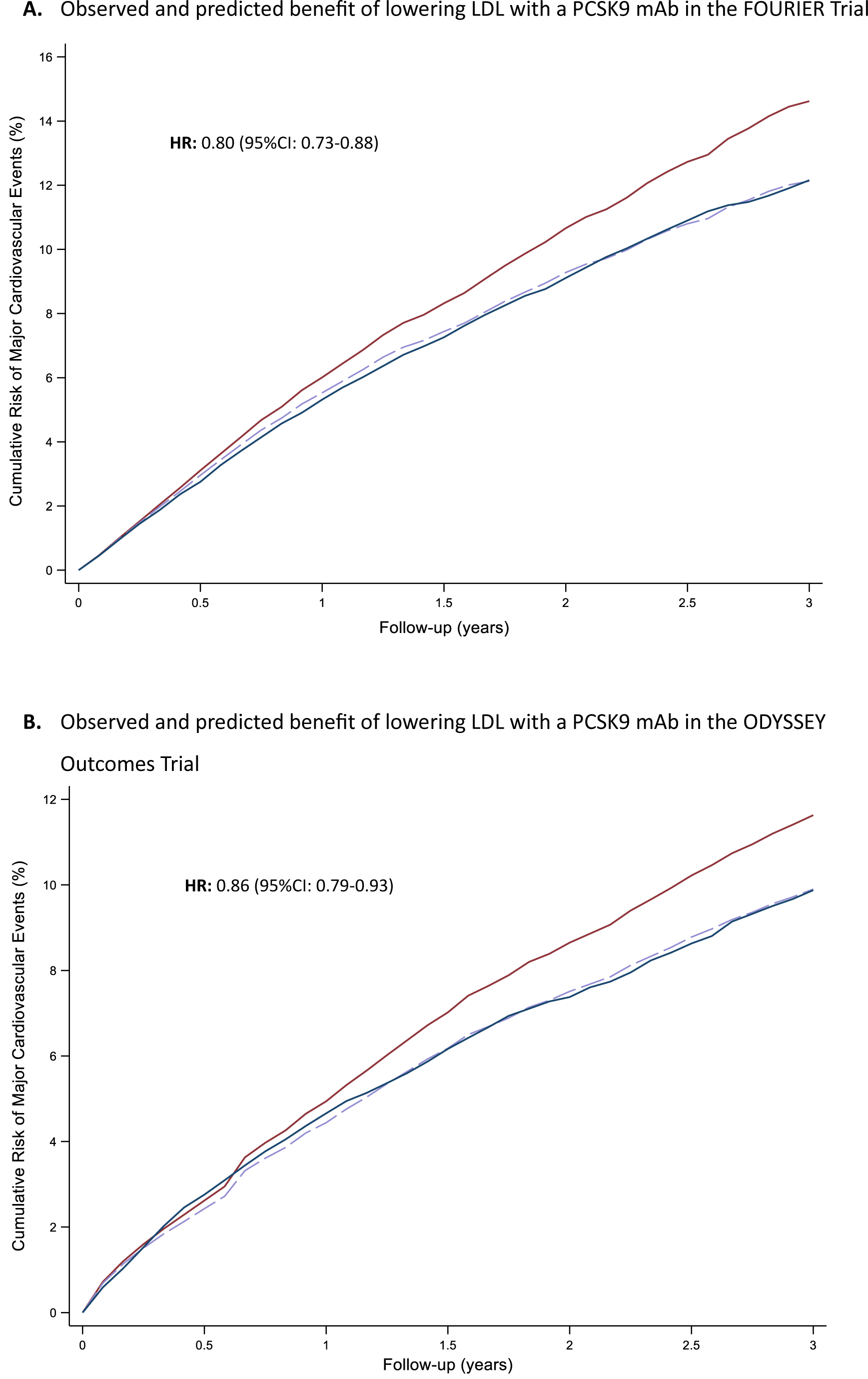
Observed and Predicted Benefit of LDL Lowering Started Later in Life on Risk of Major Cardiovascular Events. Panel A shows the cumulative rate of major cardiovascular events during a median of 2.2 years of follow-up among 27,564 participants enrolled in the FOURIER trial. Panel B shows the cumulative rate of major cardiovascular events during a median of 2.8 years of follow-up among 18,924 participants enrolled in the ODYSSEY Outcomes trial. In both panels, the red line shows the cumulative event curve for the participants who were randomized to placebo and the blue line shows the cumulative event curve for participants randomized to treatment with a monoclonal antibody directed against PCSK9. The dashed lavender line represents the cumulative event curve predicted by the causal AI algorithm for participants randomized to a PCSK9 monoclonal antibody based on the observed difference in LDL between the two treatment groups during follow-up. HR is hazard ratio, and CI is confidence interval.

Similarly, the 18,924 participants enrolled in the ODYSSEY Outcomes trial, 2,099 participants experienced a first major cardiovascular event during a median of 2.8 years of follow-up.^12^ Participants randomized to treatment with a PCSK9 mAb beginning at an average age of 58.5 years had a time-averaged 43.5 mg/dL lower LDL and a 14% lower risk of major cardiovascular events (HR: 0.86, 95%CI: 0.79-0.93, p=6.3×10^-5^). Once again, the causal AI algorithm accurately estimated the benefit of lowering due to treatment with a PCSK9 mAb during every month of follow-up during the trial, with nearly superimposable predicted and observed event curves (Figure 2B).

### ESTIMATING BENEFIT OF LDL LOWERING STARTING AT DIFFERENT AGES

We note that the PCSK9 siRNA is designed to explicitly recapitulate the LDL lowering effect of partial loss-of-function variants in the PCSK9 gene. Therefore, because the causal AI algorithm accurately estimated both the long-term benefit of lifelong lower LDL due to partial loss of function of the PCSK9 gene and the short-term benefit of lowering LDL by inhibiting PCSK9 with a mAb starting at an average age of 58.5 to 62.5 years in the randomized trials, we proceeded to the next stage of the study and used the causal AI algorithm to estimate the benefit of lowering LDL by inhibiting PCSK9 with an siRNA beginning at difference ages and under different scenarios among 1.1 million men and 1.2 million women from the UK General Practice Research Database.

We began by comparing the expected benefit of lowering LDL with a PCSK9 siRNA starting at ages 30, 40, 50, and 60 years of age. Lowering LDL by a time-averaged 36% (49.0 mg/dL or 1.3 mmol/L) with a once-yearly dose of an siRNA directed against PCSK9 is estimated to reduce the total lifetime risk of major cardiovascular events before the age of 80 years by 57% (HR: 0.43, 95%CI: 0.39-0.47) if started at age 30, by 48% (HR: 0.52, 95%CI: 0.50-0.54) if started at age 40, by 38% (HR: 0.62, 95%CI: 0.60-0.65) if started at age 50, and by 26% (HR: 0.74, 95%CI: 0.71-0.77) if started at age 60 and continued to age 80 years among men (Figure 3). A similar step-wise reduction in the total lifetime risk of major cardiovascular events before the age of 90 years would be expected among women, noting that the event curves for atherosclerotic cardiovascular events appear to be delayed by approximately a decade among women (Supplemental Figure 2).

**Figure 3:**
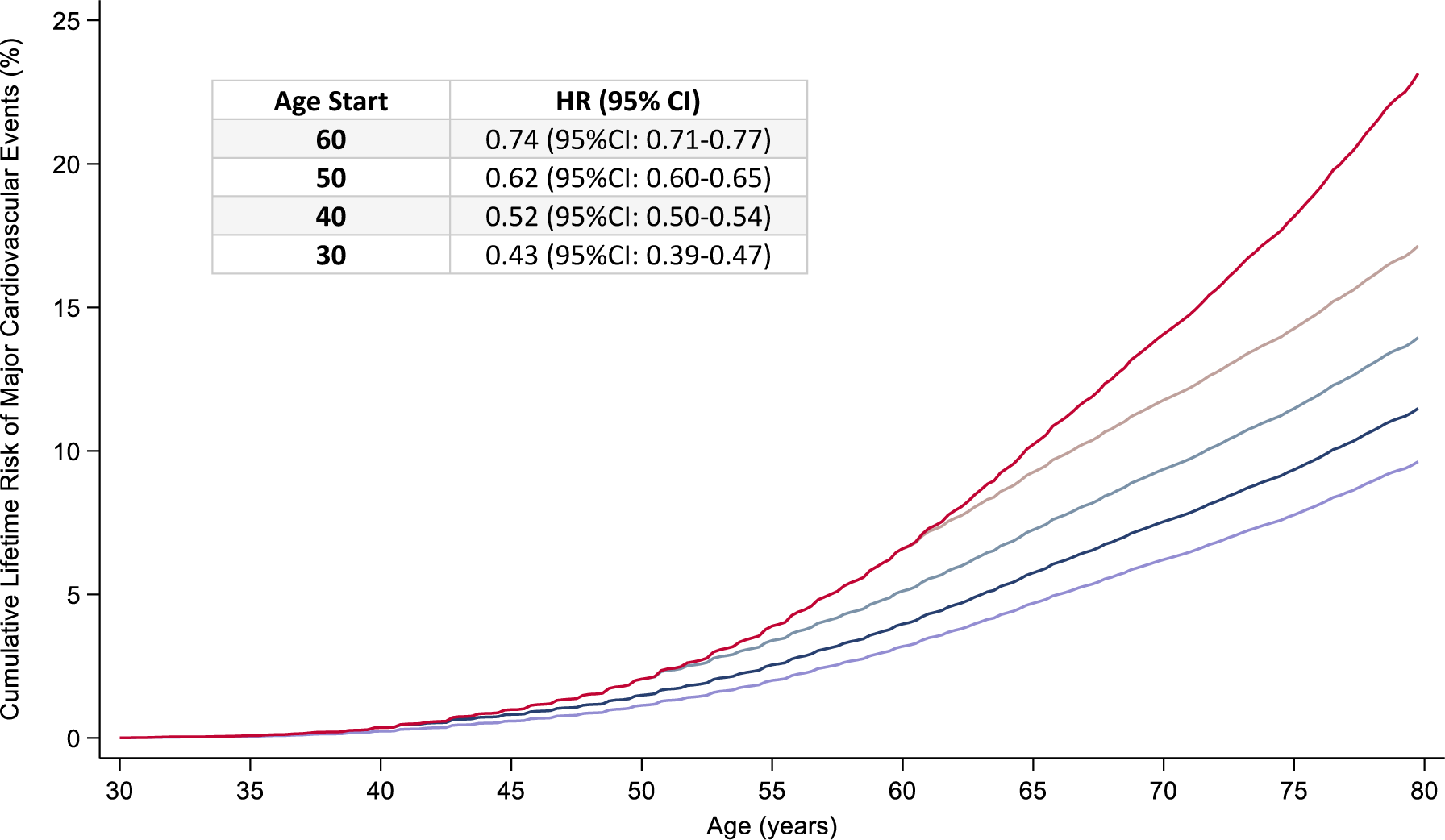
Estimated Benefit of Lowering LDL Beginning at Different Ages on the Lifetime Risk of Major Cardiovascular Events. The red line displays the observed cumulative rate of major cardiovascular events at all ages up to 80 years among 1.1 million men included in the UK General Practice Research Database. The lower lines display the estimated cumulative event curves for the same population in response to a 36% time-averaged reduction in LDL, approximately what can be achieved with a once-yearly dose of an siRNA directed against PCSK9, beginning at ages 30, 40, 50, and 60 years. HR is hazard ratio, and CI is confidence interval.

### COMPARING BENEFIT OF MODEST EARLY AND AGGRESSIVE LATER LDL LOWERING

Lowering LDL by a time-averaged 36% with a once-yearly dose of an siRNA directed against PCSK9 starting at age 40 years is estimated to reduce the total lifetime risk of major cardiovascular events more than lowering LDL by a time-averaged 52% (70.2 mg/dL or 1.8 mmol/L) with a twice-yearly dose of a PCSK9 siRNA started at age 55 years (Figure 4). Although more aggressive LDL lowering would reduce the instantaneous hazard ratio of experiencing a major cardiovascular event as compared to less aggressive LDL lowering, the absolute cumulative risk of experiencing a major cardiovascular event would remain lower at all ages among persons who started modest lowering LDL earlier as compared to more aggressive LDL lowering started later, despite achieving approximately the same total cumulative reduction in LDL before the age of 80 years (Supplemental Figures 3 and 4).

**Figure 4:**
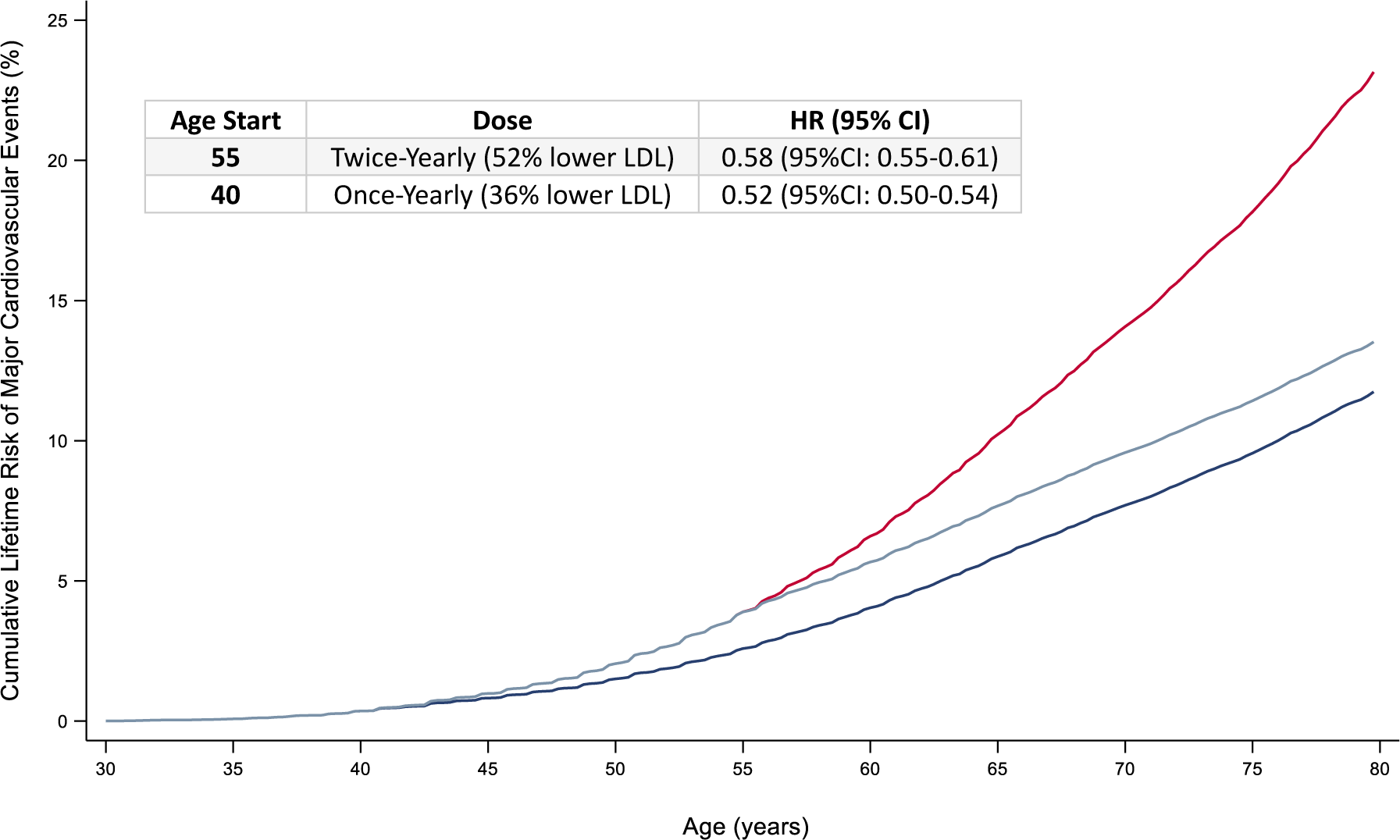
Comparing the Estimated Benefit of Moderate LDL Lowering Started Earlier with More Aggressive LDL Lowering Started Later on the Lifetime Risk of Major Cardiovascular Events. The red line displays the observed cumulative rate of major cardiovascular events at all ages up to 80 years among 1.1 million men included in the UK General Practice Research Database. The lower lines display the estimated cumulative event curves for the same population in response to a 36% time-averaged reduction in LDL (approximately what can be achieved with a once-yearly dose of an siRNA directed against PCSK9) beginning at age 40 years, and a 52% time-averaged reduction in LDL (approximately what can be achieved with twice-yearly doses of an siRNA directed against PCSK9) beginning at age 55 years. HR is hazard ratio, and CI is confidence interval.

### LEGACY BENEFIT OF EARLIER LDL LOWERING

Finally, we compared the expected benefit of the age at which LDL lowering is started on the remaining lifetime risk of major cardiovascular events among persons who survive to different ages. This analysis is designed to evaluate the potential legacy benefit lowering LDL starting early in life among persons who survive to middle age without a cardiovascular event. Lowering LDL by a time-averaged 36% with a once-yearly dose of an siRNA directed against PCSK9 is estimated to reduce the remaining lifetime risk of major cardiovascular events among persons who survive to age 55 without a cardiovascular event by 54% (HR: 0.46, 95%CI: 0.40-0.51) if started at age 30, by 47% (HR: 0.53, 95%CI: 0.50-0.57) if started at age 40, and by 34% (HR: 0.66, 95%CI: 0.61-0.71) if started at age 55 among men (Figure 5). A similar magnitude of this legacy benefit is estimated reducing the remaining lifetime risk of cardiovascular events before the age of 90 years among women (Supplemental Figure 5). The magnitude of the expected legacy benefit of starting an siRNA early can be quantified as the difference in the area under the cumulative event curves by the age at which LDL lowering is started.

**Figure 5:**
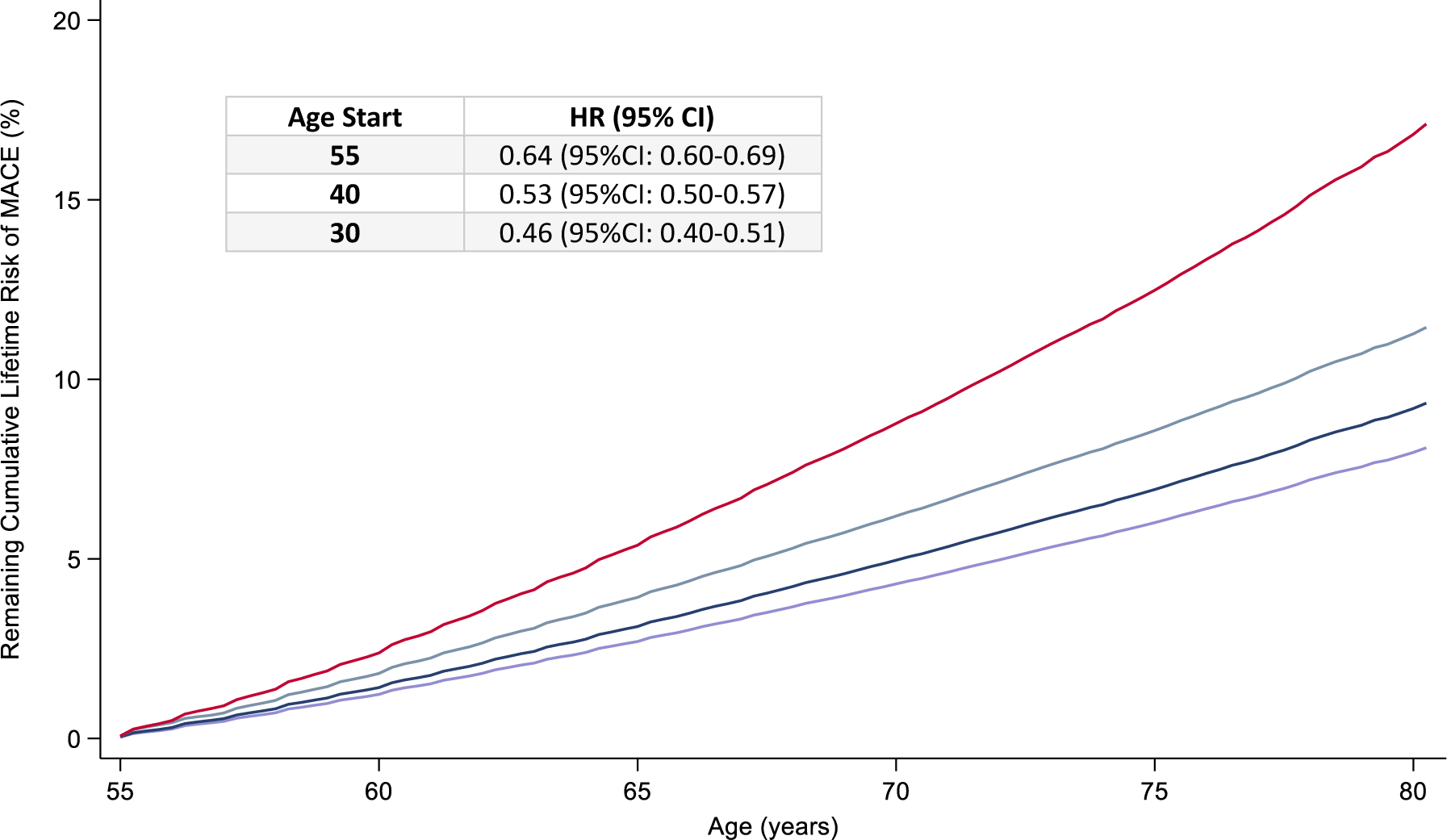
Estimating the Legacy Benefit of Early LDL lowering on the Remaining Lifetime Risk of Major Cardiovascular Events. The red line displays the observed cumulative rate of major cardiovascular events at all ages up to 80 years among the 1.1 million men included in the UK General Practice Research Database who survived to age 55 without experiencing a major cardiovascular event. The lower lines display the estimated cumulative event curves for the same population in response to a 36% time-averaged reduction in LDL, approximately what can be achieved with a once-yearly dose of an siRNA directed against PCSK9, beginning at ages 30, 40, and 55 years. HR is hazard ratio, and CI is confidence interval.

## DISCUSSION

We developed a casual AI algorithm that encodes the biological effect of LDL on the risk of atherosclerotic cardiovascular events in discrete time-units of exposure, conditional on previous exposure to take into account the size of the accumulated plaque burden at the time LDL lowering is started, to estimate the clinical benefit of lowering LDL beginning at any age and extending for any duration. We found that this causal AI algorithm accurately predicted the reduction in major cardiovascular events during every year of life among participants randomized by nature to lifelong exposure to lower LDL-C due to a partial loss-of-function variant in the PCSK9 gene; and accurately predicted the reduction in major cardiovascular events during every month of follow-up among participants randomized to treatment to lower LDL-C with a monoclonal antibody directed against PCSK9 for up to three years starting at a mean age of 61 years in two large randomized trials – with nearly superimposable observed and predicted cumulative event curves in all analyses. We then used this algorithm to estimate the clinical benefit of lowering LDL-C beginning at different ages throughout life, and found that the proportional reduction in the risk of major cardiovascular events increases with each decade earlier that LDL-C lowering is started. In addition, we found that modest early LDL lowering is associated with a greater reduction in the risk of major cardiovascular events at all ages as compared to more intense LDL lowering started later in life; and that the legacy benefit of early LDL lowering persists largely unchanged throughout life. These findings challenge the current prevention paradigm and have potentially important clinical implications for how to more effectively prevent atherosclerotic cardiovascular events.

The results of this study confirm that the benefit of lowering LDL is determined by both the magnitude and duration of LDL lowering. Each decade earlier that LDL lowering is started appears to result in a progressively greater proportional reduction in the risk of major cardiovascular events for the same reduction in LDL, presumably by slowing the progression of atherosclerosis to reduce the size of the accumulated plaque burden at any point in time. Therefore, a once-yearly dose of an siRNA directed against PCSK9, or other therapy that can produce sustained reductions in LDL over a prolonged duration of time, could potentially substantially reduce the lifetime risk of cardiovascular events by slowing the progression of atherosclerosis.^10,22^

Furthermore, modest LDL lowering started earlier appears to produce a greater benefit compared to more aggressive LDL lowering started later in life. This finding suggests that the duration of LDL lowering may be more important than the magnitude of LDL lowering for reducing the lifetime risk of major cardiovascular events. Indeed, modest early LDL lowering appears to be associated with a lower residual risk of cardiovascular events at all ages as compared to more aggressive LDL lowering started later in life. This finding implies that the residual risk of cardiovascular events during LDL lowering therapy may be due to the size of the plaque burden that accumulates before LDL lowering is started, thus providing a biological explanation for why the duration of LDL lowering may be more important than the magnitude of LDL lowering.

In addition, the benefit of earlier LDL lowering appears to persist largely unattenuated over time to create a legacy benefit. The results of this study suggest that among persons who survive to a certain age without a cardiovascular event and are treated to the same LDL level, those persons who started LDL lowering earlier have a lower residual risk of cardiovascular events at all ages as compared to persons who started LDL lowering later. Therefore, the legacy benefit of early LDL lowering can be quantified as the difference in residual risk during treatment based on when LDL lowering was started. This conclusion suggests the timing of LDL lowering is important. Earlier LDL lowering slows the progression of atherosclerosis resulting in a smaller accumulated plaque burden thus reducing the residual risk of cardiovascular events at any point in time. By contrast, starting LDL lowering later in life means the accumulated plaque burden will be much larger at the time LDL lowering is started thus resulting in a higher residual risk of cardiovascular events, and providing a biological explanation for the legacy effect of early LDL lowering.

Synthesizing these lines of evidence leads to the conclusion that the clinical benefit of lowering LDL is determined by the magnitude, duration, and timing of LDL lowering. Therefore, modest sustained LDL lowering beginning early in life may be the optimal strategy to prevent atherosclerotic cardiovascular events. This strategy should produce the greatest reduction in the lifetime risk of atherosclerotic cardiovascular events by most effectively slowing the progression of atherosclerosis and thus minimizing the residual risk of cardiovascular events at all ages. Because the casual AI algorithm accurately estimated the long-term benefit of maintaining lower LDL during every year of life among persons with a partial LOF variant in the PCSK9 gene and estimated the short-term benefit of lowering LDL starting later in life during every month of follow-up during treatment with a PCSK9 mAb, this algorithm should be able to accurately estimate the benefit of lowering LDL beginning at any age and thus can be used to provide guidance about the optimal timing and intensity of lowering LDL to prevent atherosclerotic cardiovascular events by slowing the progression of atherosclerosis.

This study has limitations. First, all analyses were restricted to persons of self-identified European ancestry. Therefore, the study should be repeated among participants of other ethnicities. However, the causal AI algorithm explicitly attempted to code the biological effect of LDL on the risk of atherosclerotic cardiovascular disease which should be the same regardless of ethnic background. Second, the optimal study design to estimate the benefit of lowering LDL with a once-yearly dose of an siRNA directed PCSK9 beginning at different ages on the lifetime risk of cardiovascular events would be a very large randomized trial with 50 years of follow-up. However, such a trial is both impractical and unfeasible. We note that an siRNA directed against PCSK9 is explicitly designed to recapitulate the LDL lowering effect of partial loss-of-function variants in the PCSK9 gene.^23^

Therefore, a randomized trial evaluating the benefit of long-term LDL lowering by inhibiting PCSK9 would be designed to recapitulate the effect of lower LDL observed among persons who inherit a partial loss-of-function variant in the PCSK9 gene. This study attempted to solve that problem by combining the randomized evidence from Mendelian randomization studies and randomized trials to encode the biological effect of lowering LDL on the risk of cardiovascular events into a causal AI algorithm to estimate the benefit of lowering LDL by inhibiting PCSK9 by any amount and extending for any duration.

In conclusion, the benefit of lowering LDL is determined by the magnitude, duration, and timing of LDL lowering. Modest sustained LDL lowering beginning in early to middle adulthood, which can be achieved with a once-yearly dose of a PCSK9 siRNA, may be the optimal strategy to prevent atherosclerotic cardiovascular events by slowing the progression of atherosclerosis.

## Data Availability

All data produced in the present study are available upon reasonable request to the authors

## REFERENCES

1. Williams, K. J. & Tabas, I. The response-to-retention hypothesis of early atherogenesis. Arterioscler Thromb Vasc Biol 15, 551–561, doi:10.1161/01.atv.15.5.551 (1995).

2. Ference, B. A. et al. Low-density lipoproteins cause atherosclerotic cardiovascular disease. 1. Evidence from genetic, epidemiologic, and clinical studies. A consensus statement from the European Atherosclerosis Society Consensus Panel. Eur Heart J 38, 2459-2472, doi:10.1093/eurheartj/ehx144 (2017).

3. Boren, J. et al. Low-density lipoproteins cause atherosclerotic cardiovascular disease: pathophysiological, genetic, and therapeutic insights: a consensus statement from the European Atherosclerosis Society Consensus Panel. Eur Heart J 41, 2313–2330, doi:10.1093/eurheartj/ehz962 (2020).

4. Ference, B. A., Graham, I., Tokgozoglu, L. & Catapano, A. L. Impact of Lipids on Cardiovascular Health: JACC Health Promotion Series. Journal of the American College of Cardiology 72, 1141–1156, doi:10.1016/j.jacc.2018.06.046 (2018).

5. Ference, B. A., Kastelein, J. J. P. & Catapano, A. L. Lipids and Lipoproteins in 2020. Jama 324, 595–596, doi:10.1001/jama.2020.5685 (2020).

6. Cohen, J. C., Boerwinkle, E., Mosley, T. H., Jr. & Hobbs, H. H. Sequence variations in PCSK9, low LDL, and protection against coronary heart disease. N Engl J Med 354, 1264–1272, doi:10.1056/NEJMoa054013 (2006).

7. Ference, B. A. et al. Effect of long-term exposure to lower low-density lipoprotein cholesterol beginning early in life on the risk of coronary heart disease: a Mendelian randomization analysis. Journal of the American College of Cardiology 60, 2631–2639, doi:10.1016/j.jacc.2012.09.017 (2012).

8. Ference, B. A. et al. Variation in PCSK9 and HMGCR and risk of cardiovascular disease and diabetes. N Engl J Med 375, 2144–2153, doi:10.1056/NEJMoa1604304 (2016).

9. Ray, K. K. et al. Effect of 1 or 2 Doses of Inclisiran on Low-Density Lipoprotein Cholesterol Levels: One-Year Follow-up of the ORION-1 Randomized Clinical Trial. JAMA Cardiol 4, 1067–1075, doi:10.1001/jamacardio.2019.3502 (2019).

10. Braunwald, E. How to live to 100 before developing clinical coronary artery disease: a suggestion. European Heart Journal 43, 249–250, doi:10.1093/eurheartj/ehab532 (2021).

11. Ference, B. A. How to use Mendelian randomization to anticipate the results of randomized trials. Eur Heart J 39, 360–362, doi:10.1093/eurheartj/ehx462 (2018).

12. Sabatine, M. S. et al. Evolocumab and clinical outcomes in patients with cardiovascular disease. N Engl J Med 376, 1713–1722, doi:10.1056/NEJMoa1615664 (2017).

13. Schwartz, G. G. et al. Alirocumab and cardiovascular outcomes after acute coronary syndrome. N Engl J Med 379, 2097–2107, doi:10.1056/NEJMoa1801174 (2018).

14. Board, J. B. S. Joint British Societies’ consensus recommendations for the prevention of cardiovascular disease (JBS3). Heart 100 Suppl 2, ii1-ii67, doi:10.1136/heartjnl-2014-305693 (2014).

15. Ray KK, et al.; ORION-10 and ORION-11 Investigators. Two Phase 3 Trials of Inclisiran in Patients with Elevated LDL Cholesterol. N Engl J Med. 382(16):1507-1519. doi: 10.1056/NEJMoa1912387. (2020).

16. Catapano, A. L., Pirillo, A. & Norata, G. D. Insights from ORION studies: focus on inclisiran safety. Cardiovasc Res, doi:10.1093/cvr/cvaa139 (2020).

17. Heart Protection Study Collaborative Group. Effects on 11-year mortality and morbidity of lowering LDL cholesterol with simvastatin for about 5 years in 20,536 high-risk individuals: a randomised controlled trial. Lancet. 378(9808):2013-2020. doi: 10.1016/S0140-6736(11)61125-2. (2011).

18. Sudlow, C. et al. UK biobank: an open access resource for identifying the causes of a wide range of complex diseases of middle and old age. PLoS Med, 12(3):e1001779, (2015).

19. Baigent, C. et al. Efficacy and safety of more intensive lowering of LDL cholesterol: a meta-analysis of data from 170,000 participants in 26 randomised trials. Lancet 376, 1670–1681, doi:10.1016/S0140-6736(10)61350-5 (2010).

20. Yusuf S, et al.; HOPE-3 Investigators. Cholesterol Lowering in Intermediate-Risk Persons without Cardiovascular Disease. N Engl J Med. 374(21):2021–31. doi: 10.1056/NEJMoa1600176. (2016).

21. O’Donoghue, M. L. et al. Long-Term Evolocumab in Patients With Established Atherosclerotic Cardiovascular Disease. Circulation 146, 1109–1119, doi:10.1161/CIRCULATIONAHA.122.061620 (2022).

22. Ray, K. K. et al. World Heart Federation Cholesterol Roadmap 2022. Global heart 17, 75, doi:10.5334/gh.1154 (2022).

23. Ference, B. A., Holmes, M. V. & Smith, G. D. Using Mendelian Randomization to Improve the Design of Randomized Trials. Cold Spring Harb Perspect Med 11, doi:10.1101/cshperspect.a040980 (2021).

